# Key features associated with opioid misuse in chronic pain: A machine learning cross-sectional study

**DOI:** 10.1101/2025.08.06.25333075

**Authors:** Dokyoung S. You, Yiyu Wang, Samsuk Kim, Mark A. Lumley, Beth D. Darnall, Sean C. Mackey, Maisa S. Ziadni

**Affiliations:** Department of Family and Community Medicine, Health Promotion Research Center, Stephenson Cancer Center, University of Oklahoma-Tulsa; Department Anesthesiology, Perioperative and Pain medicine, Stanford University School of Medicine, 1070 Arastradero Road, Suite 200, MC 5596, Palo Alto, CA 94304; Department of Psychology, Wayne State University, 5057 Woodward, Suite 7908 Detroit, Michigan 48202

**Keywords:** Prescription opioid misuse, Chronic pain, Anger, Ambivalence over emotion expression, Machine learning

## Abstract

**Importance:** Opioid misuse remains a critical public health concern, associated with increased risk of overdose, psychiatric comorbidity, and societal costs. While machine learning (ML) analyses have been used to identify features associated with opioid misuse across various clinical settings, few studies have examined both modifiable and non-modifiable features. Understanding modifiable features may inform targeted prevention and intervention strategies to reduce opioid misuse.

**Objective:** To identify key features associated with opioid misuse severity in patients with chronic pain taking long-term prescription opioids.

**Design, Setting, and Participants:** This analysis used data from clinical trials investigating a skills-based pain management intervention. Participants were 314 community-dwelling adults with chronic pain who were taking daily opioid medications (≥ 10 morphine-equivalent daily dose) for at least 3 months. Data was extracted from baseline assessments.

**Outcomes and Measures:** Opioid misuse severity was assessed using the Current Opioid Misuse Measure (COMM). Thirty-six demographic and clinical features were evaluated, including PROMIS symptom domains (e.g., pain rating, pain interference, physical function, fatigue, sleep disturbance, depression, anxiety, and anger) as well as emotional ambivalence, pain catastrophizing, trauma exposure, and substance use.

**Results:** Among the seven ML algorithms (Random Forest, XGBoost, Support Vector Regression, LASSO regression, Ridge Regression, Elastic Net, and Multilayer Perceptron), the Elastic Net model demonstrated the strongest performance, yielding the highest correlation with COMM scores (mean *r* = 0.61, 95% CI [0.40, 0.73]) and the lowest root mean square error (mean RMSE = 4.16, 95% CI [3.41, 4.80]). Feature ablation analysis identified anger (Δ*r* = 0.053), emotional ambivalence (Δ*r* = 0.022), pain catastrophizing (Δ*r* = 0.008), and fatigue (Δ*r* = 0.002) as the most influential features associated with the COMM scores. Shapley Additive exPlanations (SHAP) analysis confirmed that higher levels of these key features were associated with higher COMM scores.

**Conclusions and Relevance:** Emotional factors, particularly anger, emerged as key features associated with the severity of opioid misuse. These findings suggest that interventions targeting emotion regulation, especially anger management, may reduce opioid misuse among individuals receiving long-term opioid therapy for chronic pain.

## Introduction

Chronic pain is a debilitating condition that affects 50.2 million (20.5%) adults in the United States (U.S.).^1^ Musculoskeletal disorders such as chronic back pain and arthritis are the leading causes of disability in the U.S.^2,3^ To manage chronic pain, 29.3% of affected individuals use prescription opioid.^4^ Opioids are particularly utilized by those with worse health status, including higher levels of pain, pain interference, fatigue, sleep disturbance, and depression.^5^ Consistent with this pattern, its utilization is higher among those with activity-limiting pain,^6^ with 41.2% reporting prescription opioid use.^4^

Opioid medications are generally used as prescribed; however, they carry a risk of misuse. The prevalence of opioid misuse is estimated to be between 21% and 29% among individuals using opioids for pain management.^7^ Misuse refers to taking opioids in a manner other than prescribed, such as using higher doses, more frequently, or for a longer duration than directed.^8^ It also includes using opioids for non-medical reasons, taking someone else’s prescription, continuing use despite negative consequences, or combining opioids with central nervous system depressants (e.g., benzodiazepines, alcohol).^9^ Opioid misuse is a significant health concern, as it increases the risk of overdose and is associated with greater emotional distress.^5,10^ Furthermore, it imposes a substantial financial burden. Individuals with opioid misuse spend $15,000 more on healthcare annually compared to those without it, and the social cost is estimated at $50 billion per year.^11^

To address opioid misuse, machine learning (ML) has been used to identify risk factors across various healthcare settings. ML models can capture complex, non-linear interactions without strict parametric assumptions. Due to this flexibility, ML models are better suited in detecting risk factors for multifaceted conditions like opioid misuse. One ML study analyzing electronic health records (EHRs) from intensive care units (ICU) identified predictors of opioid misuse, including: a) ICD-9 codes related to drug withdrawal, chronic hepatitis, posttraumatic stress disorder (PTSD), tobacco use disorder, dysthymic disorder, unspecified drug abuse, cocaine abuse, and other chronic pain; and b) certain prescription medications such as methadone, nicotine patch, diazepam, zolpidem, quetiapine, and clonazepam.^12^ Another ML study analyzing EHRs of surgical patients identified pre-operative opioid use, depression, and antidepressant use as predictors of prolong opioid misuse after surgery.^13^ However, no study has applied ML to community-dwelling adults with chronic pain on long-term opioids. Moreover, prior ML studies have rarely incorporated modifiable psychological risk factors, which may inform behavioral treatment targets.

Traditional regression analyses have identified psychological risk factors for opioid misuse in chronic pain populations, such as pain catastrophizing^10,14-16^ and emotion dysregulation.^17,18^ Pain catastrophizing refers to a negative cognitive and emotional response to pain.^19^ Emotion dysregulation refers to the inability to regulate emotional reactions in adaptive ways.^20^ Specifically, it involves difficulties with awareness, understanding, and accepting emotions, as well as engaging in a goal-directed behavior or inhibit inappropriate or impulsive behaviors when experiencing negative emotions.^20^ Recently, ambivalence over emotion expression (hereafter emotional ambivalence), a form of emotion dysregulation, has been identified as a factor associated with opioid misuse in individuals with chronic pain.^21^

The current cross-sectional study applied an ML approach to examine both modifiable and non-modifiable factors associated with opioid misuse severity among adults with chronic pain receiving long-term opioid therapy. The primary aim was to identify the most influential features, with particular interest in pain catastrophizing and emotional ambivalence.

## Methods

### Study Design

Stanford’s Institutional Review Board approved this study procedure. Participants were recruited for three clinical trials, investigating the effect of a single-session, pain self-management skills class. Recruitment began in 2019 and ended in 2024. The current study analyzed the baseline data from different phases of clinical trials, which included: participants enrolled in the in-person version of the trial prior to COVID (N=46), single-arm Zoom-delivered version during COVID (N=55), and the final Zoom-delivered RCT (N=213). Inclusion and exclusion criteria were consistent across all three phases. Participants were eligible if they: a) were at least 18 years old, b) had chronic non-cancer pain (at least 6-month duration), c) had been using prescription opioids (≥ 10 morphine-equivalent daily dose: MEDD) for ≥ 3 months, d) were fluent in English, and e) had access to an electronic device to complete web-based surveys and attend a zoom-delivered class. Exclusion criteria included: a) ongoing litigation or disability claim, b) concurrent participation in another pain self-management skills class, c) inability to complete any of the study required tasks (e.g., cognitive impairment), and d) active suicidality. Eligible participants received the electronic informed consent, which was administered via the REDCap, Health Insurance Portability and Accountability Act (HIPAA)–compliant platform.^22^ Once signed the informed consent, baseline surveys were administered through REDCap.

## Measures

### Descriptive demographic and clinical variables

Demographic information, including age, gender, race/ethnicity, income, marital status, and employment status, was collected. Additionally, data on pain diagnoses, current disability status, mental health diagnoses, and history of psychological treatment were gathered. For pain and mental health diagnoses, participants responded to yes/no questions and, if endorsed, they were prompted to specify their conditions in a free-text format.

### Outcome variable: Opioid misuse

The severity of opioid misuse was assessed using the 17-item Current Opioid Misuse Measure (COMM), which has demonstrated excellent internal consistency (α= 0.86^23^ and 0.83^24^) and reliability (ICC = 0.86).^24^ The cutoff sore of 9 has been validated to identify problematic opioid misuse.^24^ As expected, our sample’s COMM scores were rightly skewed (Figure S1).

### Independent variables

Thirty-six demographic and clinical variables were examined (Table 1.1 and 1.2). The correlation matrix illustrates the relationships among demographic and clinical variables (Figure S1). Income and education were dummy coded such that higher values would reflect greater income and higher levels of education (see Table 1.1 note for coding details). To improve model efficiency and reduce overfitting, employment status, marital status, race, and disability were recoded as binary variables: “employed vs unemployed,” “married vs non-married,” “white vs non-white,” and “disability vs no disability.” Binary coding reduced the number of predictors and addressed sparse category representation, improving model convergence.

**Table 1.1.** Demographic characteristics (n = 314)

| Variables |  | M | SD | Range |
| --- | --- | --- | --- | --- |
| **Age** | (years) | 54.4 | 11.9 | 19 - 86 |
|  |  | N | (%) |  |
| Gender | **Female** | 241 | 76.8 |  |
|  | Male | 71 | 22.6 |  |
|  | Other (including decline to answer) | 2 | 0.6 |  |
| Ethnicity | Non-Hispanic | 297 | 94.6 |  |
|  | **Hispanic** | 17 | 5.4 |  |
| Race | **White/Caucasian** | 282 | 89.8 |  |
|  | Black/African | 9 | 2.9 |  |
|  | Asian | 5 | 1.6 |  |
|  | Native Hawaiian or Pacific Islander | 1 | 0.3 |  |
|  | American Indian or Alaska Native | 3 | 1.0 |  |
|  | More than one | 10 | 3.2 |  |
|  | Other or unknown | 4 | 1.3 |  |
| Marital Status | **Married** | 168 | 53.5 |  |
|  | Partnered and living together | 22 | 7.0 |  |
|  | In a relationship but not living together | 10 | 3.2 |  |
|  | Divorced | 59 | 18.8 |  |
|  | Separated | 11 | 3.5 |  |
|  | Widowed | 13 | 4.1 |  |
|  | Never married | 31 | 9.9 |  |
| Employment status* | **Employed** | 69 | 22.0 |  |
|  | Not currently employed | 18 | 5.7 |  |
|  | Temporarily laid off/sick leave or maternity leave | 3 | 1.0 |  |
|  | Disabled or not working due to pain | 178 | 54.8 |  |
|  | Retired | 74 | 23.6 |  |
|  | Other | 15 | 4.8 |  |
|  |  | M | SD | Range |
| **Income** |  | 4.6 | 2.2 | 1 - 8 |
| **Education** |  | 6.6 | 1.4 | 3 - 10 |

Note: Income brackets were 1, Less than \$10,000; 2, <\$25,000; 3, \$25,000-\$44,999; 4, \$45,000-\$64,999; 5, \$65,000-\$84,999; 6, \$85,000-\$104,999; 7, \$105,000-\$124,999; 8, ≥\$125,000. Education brackets were: 1, Grade School/Elementary School (kindergarten through 6th grade); 2, Middle School (through 8th grade); 3, Some High School; 4, High School Diploma or GED; 5, Some College, No Degree; 6, Associate's Degree or Vocational Certificate; 7, Bachelor's Degree (e.g., BA, AB, BS, BBA); 8, Master's Degree (e.g., MA, MS, MEng, MEd, MBA); 9, Professional School Degree (e.g., MD, DDS, DVM, JD); 10, Doctoral Degree (e.g., PhD, EdD). \*The total exceeds 314 because multiple selections were allowed. Variables shown in bold were included as binary variables in the ML model.

**Table 1.2.** Clinical characteristics (n = 314) ^*^Descriptives are calculated before imputation.

| Variables | N | (%) |  |
| --- | --- | --- | --- |
| **Duration of pain** |  |  |  |
| 6 – less than 12 months | 1 | 0.3 |  |
| 1 – 5 years | 24 | 7.6 |  |
| > 5 years | 289 | 92.0 |  |
| Received diagnosis for chronic pain? (Yes) | 231 | 73.6 |  |
| **Currently on disability? (Yes)** | 149 | 47.5 |  |
| COMM positive (total scores of $\geq 9$) | 66 | 27.7 | |
| DSM 5 OUD symptom positive (total score of $\geq 2$) | 51 | 16.2 | |
| Drinking alcohol? (Yes) | 149 | 47.5 |  |
| AUDIT positive (total scores of $\geq 8$) | 2 | 0.6 | |
| Use drug for non-medical reasons? (Yes) | 38 | 12.1 |  |
| Received mental health diagnosis? (Yes) | 176 | 56.1 |  |
| Receive psychological treatment? (Yes) | 148 | 47.1 |  |
|  | M | SD | Range |
| **PROMIS-Pain Intensity (0-10 scale): Worst** | 8.6 | 1.3 | 3 – 10 |
| **Average** | 5.3 | 1.6 | 2 – 10 |
| **Current** | 5.5 | 1.9 | 0 – 10 |
| **Duration of opioid medication (years)** | 12.5 | 8.1 | 0 – 55 |
| **COMM total scores** | 6.8 | 5.2 | 0 – 33 |
| **AUDIT total scores** | 1.0 | 1.6 | 0 – 16 |
| **DAST total scores** | 0.3 | 0.3 | 0 – 4 |
| **CTQ total scores** | 47.8 | 17.7 | 25 – 119 |
| **Emotional abuse** | 11.4 | 5.2 | 5 – 25 |
| **Physical Abuse** | 8.3 | 4.5 | 5 – 25 |
| **Emotional Neglect** | 11.9 | 4.6 | 5 – 25 |
| **Physical Neglect** | 7.9 | 3.3 | 5 – 25 |
| **Sexual Abuse** | 7.7 | 4.7 | 5 – 25 |
| **PROMIS (T-scores)** |  |  |  |
| **Global Physical Health** | 36.0 | 6.7 | 19.7 – 60.3 |
| **Global Mental Health** | 41.3 | 8.7 | 21.3 – 67.6 |
| **Pain Interference** | 64.4 | 6.8 | 40.7 – 77.0 |
| **Physical Function** | 35.9 | 6.6 | 20.9 – 59.7 |
| **Fatigue** | 60.4 | 9.3 | 33.1 – 77.7 |
| **Sleep Disturbance** | 57.3 | 8.7 | 30.5 – 77.5 |
| **Depression** | 53.4 | 9.6 | 38.2 – 81.1 |
| **Anxiety** | 53.8 | 9.6 | 37.1 – 82.8 |
| **Anger** | 49.2 | 8.5 | 32.9 – 79.6 |
| **PCS total scores** | 16.9 | 11.5 | 0 – 52 |
| **Helplessness** | 7.7 | 5.5 | 0 – 24 |
| **Rumination** | 6.1 | 4.4 | 0 – 16 |
| **Magnification** | 3.1 | 2.6 | 0 – 12 |
| **AEQ total scores** | 31.7 | 11.2 | 14 – 69 |

Note: Variables shown in bold were included in the ML model.

The following measures were used to assess clinical variables. Alcohol-related problems were assessed using the 10-item **<u>Alcohol Use Disorders Identification Test</u>** (AUDIT; total score range 0 to 40), with scores of ≥ 8 indicating hazardous or harmful drinking. Drug-related problems were measured with the 10-item **<u>Drug Abuse Screening Test</u>** (DAST-10; total score range 0-10), where higher total scores indicate more problems with drug use. We also administered the **<u>DSM-5 OUD symptom checklist</u>**^25^ to evaluate OUD severity; however, this variable was excluded from the ML analysis due to predictor-outcome overlap. Additionally, the **<u>Childhood Trauma Questionnaire (CTQ)</u>** was used to assess childhood maltreatment across five domains. The CTQ consists of 28 items that evaluate the frequency of specific traumatic experiences during childhood. Total scores range from 25 to 125, with higher scores indicating greater levels of reported childhood trauma. The CTQ includes five subscales: emotional abuse, physical abuse, sexual abuse, emotional neglect, and physical neglect. Each subscale contains five items rated on a 5-point Likert scale, with total scores for each domain ranging from 5 to 25 and higher scores indicating more severe maltreatment. The average CTQ total score among adults is 32 (SD = 9 and 11 for male and female, respectively).^26^ The **<u>Patient-Reported Outcomes Measurement Information System</u>** (PROMIS) short-forms were also used to assess global physical and mental health,^27^ as well as specific health domains, including pain intensity, pain interference (8a),^28^ physical function (8a), fatigue (8a), sleep disturbance (8a), depression (8a), anxiety (8a), and anger (8a) over the past seven days. T-scores were calculated using the web-based scoring system (www.assessmentcenter.net/), with a mean of 50 (SD = 10). Higher scores indicate better global health or greater symptom severity or function, depending on the domain. The 13-item **<u>Pain Catastrophizing Scale</u>** (PCS; total scores 0-52) was used to assess the degree of pain catastrophizing, consisting of perceived helpless about, ruminating on, and enhanced threat appraisal of pain. Higher scores indicate more catastrophic thinking about pain.^19^ Helplessness (6 items), rumination (4 items), and magnification (3 items) subscale scores range from 0 to 24, 0 to 16, and 0 to 12, respectively. The 14-item **<u>Ambivalence over Emotion Expression Questionnaire (AEQ)</u>** was used to measure degree of discomfort and uncertainty when expressing their negative emotions, reflecting their internal conflict between wanting to express and suppress these emotions.^21^ Each item is rated on a 1-5 scale and total summed scores range from 14 to 70, with higher scores indicating greater ambivalence about expressing emotions.

## Missing values

Most missing data were found in the DAST, CTQ and AEQ such as 89 (28%), 86 (27%), and 25 (8%) cases, respectively. The other scores had less than 2% missing data. The higher missing rates in the DAST and CTQ may reflect the sensitive nature of assessing substance use problems and childhood trauma. All missing values were replaced using multivariate imputation to preserve statistical power and minimize potential bias.

## Analysis: hyperparameters, model training, cross validation, and interpretation

For model training, we conducted a suite of ML analyses to examine features associated with COMM scores. Seven ML models were evaluated: Random Forest, XGBoost (XGB), Support Vector Regression (SVR), LASSO regression, Ridge Regression, Elastic Net, and Multilayer Perceptron (MLP; Figure 1). Hyperparameters for each model were optimized using a grid search procedure with five-fold cross validation, implemented via Scikit-learn’s GridSearchCV.^29^ The best hyperparameters were determined based on the highest average cross-validated *r*^2^ value.

**Figure 1.**
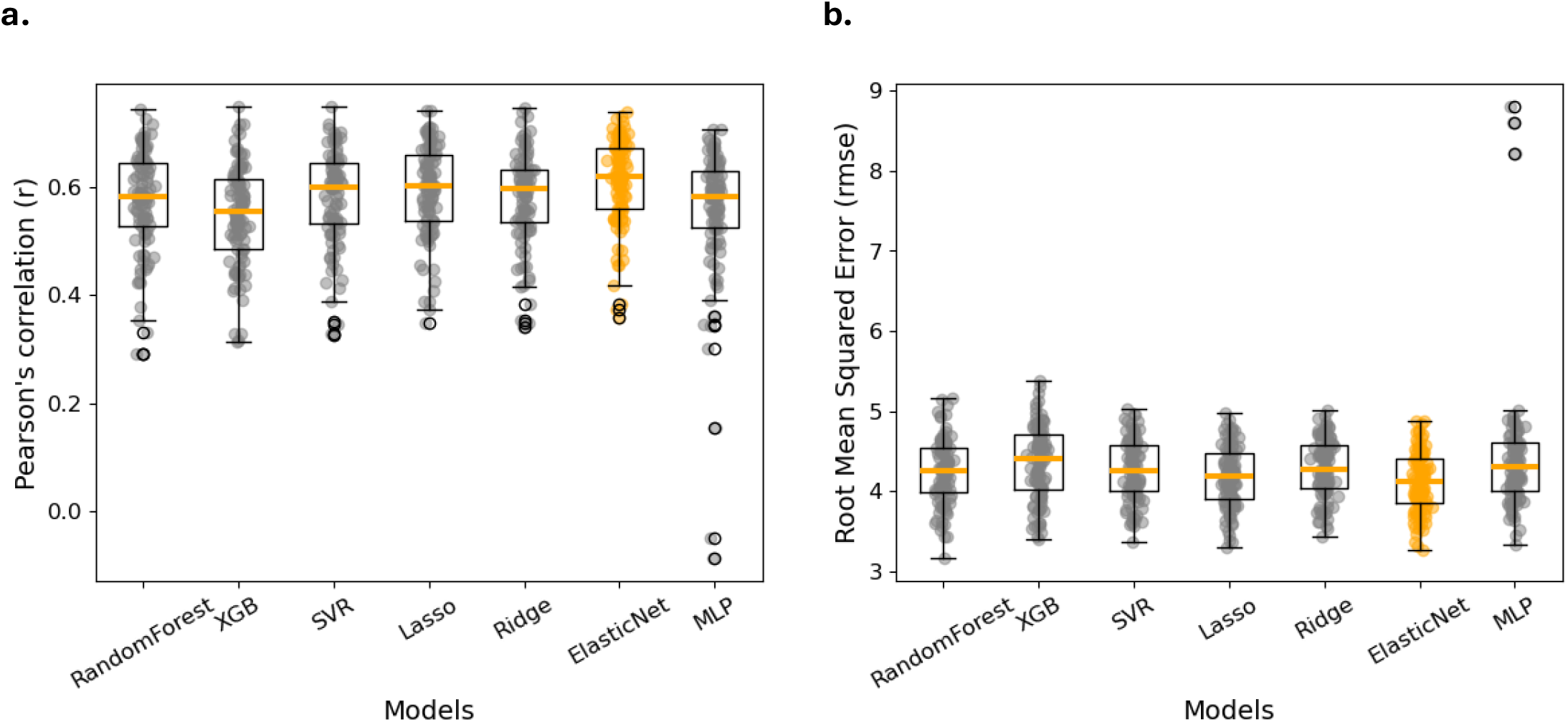
Model performance comparison. a) The distribution of the correlation coefficient (r) from the 100-fold cross validation. The Elastic Net Regression (orange) has the highest median r score (*r* = 0.61), and b) the lowest mean RMSE (RMSE = 4.12).

After hyperparameter selection, model performance was evaluated on the testing dataset to evaluate how well each trained model generalized to unseen data in predicting COMM scores. We used a 100-iteration of Monte Carlo cross-validation.^30,31^ For each iteration, the data were randomly split into a training set (80%) and a testing set (20%). The best performance model was selected based on the highest cross validated mean correlation coefficient value (*r*) (Figure 1a) and the lowest cross validated mean root mean squared error (RMSE; Figure 1b).

Following model selection, we conducted a feature ablation analysis to evaluate the relative contribution of each feature to the model performance. This involved systematically replacing the values of each independent variable with zero (i.e., “ablation”) and reapplying the model with the same 100-iteration Monte Carlo cross-validation. We then calculated the change in the model’s average correlation coefficient (*r*) compared to the original model containing all predictors. This change, referred to as Δ*r*, reflects the reduction in model performance caused by removing a specific feature. A larger Δ*r* indicates a greater decrease in model performance, suggesting that the removed feature has a greater impact on opioid misuse severity.

Next, we applied SHapley Additive exPlanations (SHAP) to the best-performing model to examine the contribution of individual features. Specifically, we examined a) the most influential features identified in a feature ablation and b) the direction of their relationship with the outcome.^32^ SHAP values quantify each feature’ contribution to the model’s prediction. A positive SHAP value associated with higher feature values indicates a positive relationship between the feature and the outcome. A wider spread of SHAP values for a given feature suggests a stronger influence on the model’s predicted outcome scores.

### Sample Size Justification

Learning curve analysis^33^ (Supplementary Figure S2) has shown that the validation score reaches plateau when training sample size reaches around 125, suggesting that the model performance is unlikely to improve by adding more data beyond this point. Therefore, our sample size of 314 would have sufficient power to detect meaningful patterns in the data.

### Study Sample

A total of 314 participants completed the baseline survey. The demographic characteristics are summarized in Table 1.1. Regarding clinical characteristics (Table 1.2), the majority (92.0%) reported that their pain duration was more than 5 years. Among those who reported their pain diagnoses (73.6%), the mean number of pain diagnosis was 2.3 (SD = 1.6, range 1 – 8) and the three most common diagnoses were fibromyalgia (19.0%), arthritis (17.7%), and chronic pain syndrome (13.0%, Supplement Table 1). The mean opioid duration was 12 years, 66 participants (27.7%) were screened positive for problematic misuse (i.e., COMM score of ≥ 9), and 51 participants (16.2%) met the diagnostic criteria for OUD. Notably, all participants were taking opioids, but moderate to severe OUD was rare (2.8%). Similarly, alcohol use was common (47.5%), but hazardous or harmful drinking was rare (0.6%). Drug use for non-medical reasons was reported by 12.1% and the most common drug was cannabis (11.5%, Supplement Table 2). Of 176 participants with reported mental health diagnoses, the average number of diagnoses was 1.7 (SD = 0.9). Depression was most common (62.5%), followed by anxiety (42.1%) and PTSD (15.9%; Supplement Table 3). About 47.1% reported receiving psychological treatments.

## Results

Among the seven models, the Elastic Net model demonstrated the strongest performance, yielding the highest mean correlation coefficient *r* of 0.61 (95% CI: 0.40, 0.73, Figure 1a) and the lowest mean RMSE of 4.12 (95% CI: 3.44, 4.85, Figure1b).

## Feature Interpretation

We performed two analyses to interpret model features. First, we conducted a feature ablation analysis to assess the relative contribution of each feature to the Elastic Net model. Because the Elastic Net uses regularization that assigns zero weights to less important features, we only conducted the feature ablation analysis on non-zero weighted features (Figure 2a). Top four features that made the most impact to the model were the PROMIS Anger T-scores (Δ*r*, = 0.053), AEQ total scores (Δ*r*, = 0.022), PCS total scores (Δ*r*, = 0.008), and the PROMIS Fatigue T-scores (Δ*r*, = 0.002). These results suggest that emotional factors such as anger and emotional ambivalence are important features associated with opioid misuse severity.

**Figure 2.**
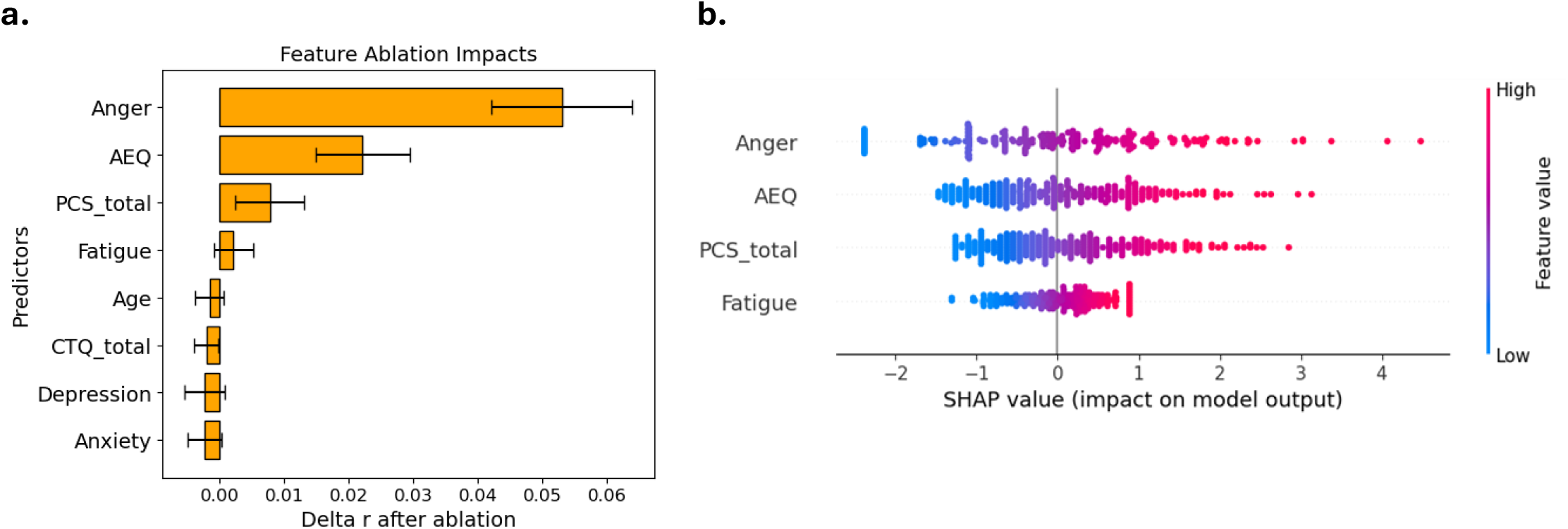
Feature Interpretation. (a)Feature ablation analysis. Only features with a non-zero delta are visualized. Given the inherent sparse regularization in Elastic Net Regression, many features were assigned zero model weights, which consequently do not affect model performance and result in zero delta r values during feature ablation. (b) SHAP summary plot of top four predictors from the feature ablation analysis. The SHAP values on the x-axis indicated the corresponding feature’s influence on the prediction. Features are color-coded from blue (low feature value) to red (high feature value). The SHAP plot provided complementary information to the feature ablation analysis by highlighting the relationship between feature values and their contributions to predicting opioid misuse.

Secondly, the SHAP model, which identifies the additive impacts of each feature, provided complementary support to the feature ablation analysis. Features including the PROMIS-Anger T-scores, AEQ scores, PCS total scores, PROMIS-Fatigue T-scores show significant impact on predicted COMM total scores (Figure 2b). Higher values of anger, AEQ, PCS, and fatigue scores (in red, Figure 2b) were associated with more positive SHAP values, indicating that greater values of these features were associated with greater severity of opioid misuse. Both feature analyses highlight the important role of emotion factors in opioid misuse.

## Discussion

The current study used an ML approach to analyze baseline data from community-dwelling adults with chronic pain who were receiving long-term opioid therapy and enrolled in clinical trials of a pain self-management skills class. Our ML model identified that anger, emotional ambivalence, pain catastrophizing, and fatigue were the most important features associated with opioid misuse severity.

Our ML model identified anger and emotional ambivalence as key features associated with opioid misuse severity. These findings are consistent with prior ML studies conducted in ICU and surgical populations, which also highlighted emotional factors as important predictors of opioid misuse.^12,13^ However, the specific emotional features associated with opioid misuse differed between the current study and previous studies. Specifically, previous ML studies based on EHRs identified depression and PTSD as primary predictors^12,13^ whereas our model identified anger as a primary factor. Notably, a prior stepwise regression analysis using data from a tertiary pain clinic sample found anxiety, anger, and depression, in that order, to be significant predictors of opioid misuse.^5^ One explanation for this discrepancy may be that anger is not routinely assessed or documented (ICD R45.4) in many clinical settings and there are no FDA-approved medications for anger. As a result, anger often goes undocumented in EHRs, limiting its detection in EHR-based research. To address this limitation, our learning healthcare system (https://choir.stanford.edu/) routinely assesses symptoms of depression, anxiety, and anger in all patients seen in a tertiary pain clinic. This comprehensive symptom tracking, in combination with an ML approach, may improve the identification of key emotional risk factors for opioid misuse in chronic pain populations.^5^

Pain catastrophizing and emotional ambivalence were of a particular interest in this current study because cumulative studies have found pain catastrophizing as a significant predictor for opioid misuse^14,15,34-36^ and a recent study identified emotional ambivalence as an additional predictor even when controlling for the known predictors like emotional distress and pain catastrophizing.^21^ Our findings extend this literature by demonstrating, through a ML approach, that both emotional ambivalence and pain catastrophizing are among the key features associated with opioid misuse severity in the chronic pain population. Besides emotional factors, fatigue was a significant feature. Fatigue is a known side effect of opioids^37^ and is positively correlated with opioid misuse severity in individuals with chronic pain.^5,10,38,39^ Future study should examine whether reducing anger, emotional ambivalence, pain catastrophizing, and fatigue would reduce opioid misuse risk.

## Limitations

Several limitations of the current study should be acknowledged. First, the cross-sectional design precludes any conclusions about causal relationship between emotional factors and opioid misuse. Second, to enhance generalizability and reduce the risk of overfitting, we employed multiple ML models and reported the final performance conservatively, based on the mean outcome from a rigorous 100-fold cross validation procedure. Future work should test the model in independent samples to evaluate its generalizability. Third, the high rates of missing data on the DAST and CTQ may have limited the model’s ability to identify substance use problems and childhood trauma as important features. Therefore, these potentially relevant factors may have been underrepresented in the final model. Finally, the sample size may have limited our ability to test more complex predictive algorithms that might enhance performance. To balance model complexity and interpretability, we prioritized model interpretability that allowed for the identification of modifiable psychological factors, thereby informing potential treatment targets for opioid misuse.

## Conclusions

The current study is the first to apply ML approach to identify key features associated with the severity of opioid misuse among community-dwelling U.S. adults with chronic pain receiving long-term opioid therapy. Consistent with previous studies, our findings underscore negative emotions as key features associated with opioid misuse in this population. Among negative emotions, anger and emotional ambivalence emerged as particularly relevant. Intervention targeting anger regulation and adaptive emotional expression may reduce the risk of opioid misuse. Building on these findings, future large-scale studies are needed to develop and validate ML-based models that can identify individuals with problematic opioid misuse. Such models could support accurate identification of opioid misuse, enable targeted intervention, and promote safer opioid prescribing practices in clinical settings.

## Data Availability

All data produced in the present study are available upon reasonable request to the authors.

## Data Sharing Statement

The data sets generated and analyzed during this study are available upon study team approval of written requests with analytic plan included in the request.

