## Supplemental_Tables_Figures for "Key features associated with opioid misuse in chronic pain: A machine learning cross-sectional study"

Supplement Table 1 The most frequently reported pain diagnosis

| Pain diagnosis | Frequency | (%) |
|---|---|---|
| 1. Fibromyalgia | 44 | 19.0 |
| 2. Arthritis | 41 | 17.7 |
| 3. Chronic pain syndrome | 30 | 13.0 |
| 4. Chronic back pain | 24 | 10.4 |
| 5. Degenerative disc disease | 21 | 9.1 |
| 6. Complex Regional Pain Syndrome | 12 | 5.2 |
| 7. Failed back syndrome | 9 | 3.9 |
| 8. Neuropathy | 7 | 3.0 |
| 9. Spinal stenosis | 6 | 2.6 |
| 10. Migraine | 5 | 2.2 |
| 11. Scoliosis | 4 | 1.7 |
| 12. Interstitial cystitis | 3 | 1.3 |
| 13. Myofascial pain syndrome | 3 | 1.3 |
| 14. Radiculopathy | 3 | 1.3 |
| 15. Spondylolisthesis | 2 | 0.9 |
| 16. Facet disease | 2 | 0.9 |
| 17. Cervical dystonia | 2 | 0.9 |
| 18. Central sensitization | 2 | 0.9 |

Note: % were calculated out of a total of 231 reported their pain diagnosis.

Supplement Table 2 Reported drug use for non-medical reasons

| Drug use | Frequency | (%) |
|---|---|---|
| Cannabis (marijuana, hashish) | 36 | 11.5 |
| Cocaine | 3 | 1.0 |
| Hallucinogens (LCD) | 3 | 1.0 |
| Narcotics (heroin, opiates) | 3 | 1.0 |
| Barbiturates (Ambien, Luminal) | 2 | 0.6 |
| Stimulants (speed, meth) | 2 | 0.6 |
| Tranquilizers (Valium, Xanax) | 1 | 0.3 |
| Solvents (Paint thinner) | 0 | 0.0 |

Note: out of a total of 314

Supplement 3 Reported mental health diagnosis

| **Mental Health Diagnosis** | **Frequency** | (%) |
|---|---|---|
| Depression disorders | 110 | 62.5 |
| Anxiety disorders | 74 | 42.1 |
| PTSD | 28 | 15.9 |
| Bipolar disorders | 9 | 5.1 |
| OCD | 7 | 4.0 |
| ADHD | 5 | 2.8 |
| Panic Disorder | 4 | 2.3 |
| Autism | 2 | 1.1 |
| Insomnia | 2 | 1.1 |
| Postpartum depression | 1 | 0.6 |
| Chemical imbalance | 1 | 0.6 |
| Agoraphobia | 1 | 0.6 |
| Dysthymia | 1 | 0.6 |
| Eating disorder | 1 | 0.6 |
| Social Anxiety | 1 | 0.6 |

Note: % was calculated out of a total of 176 with mental health diagnosis.

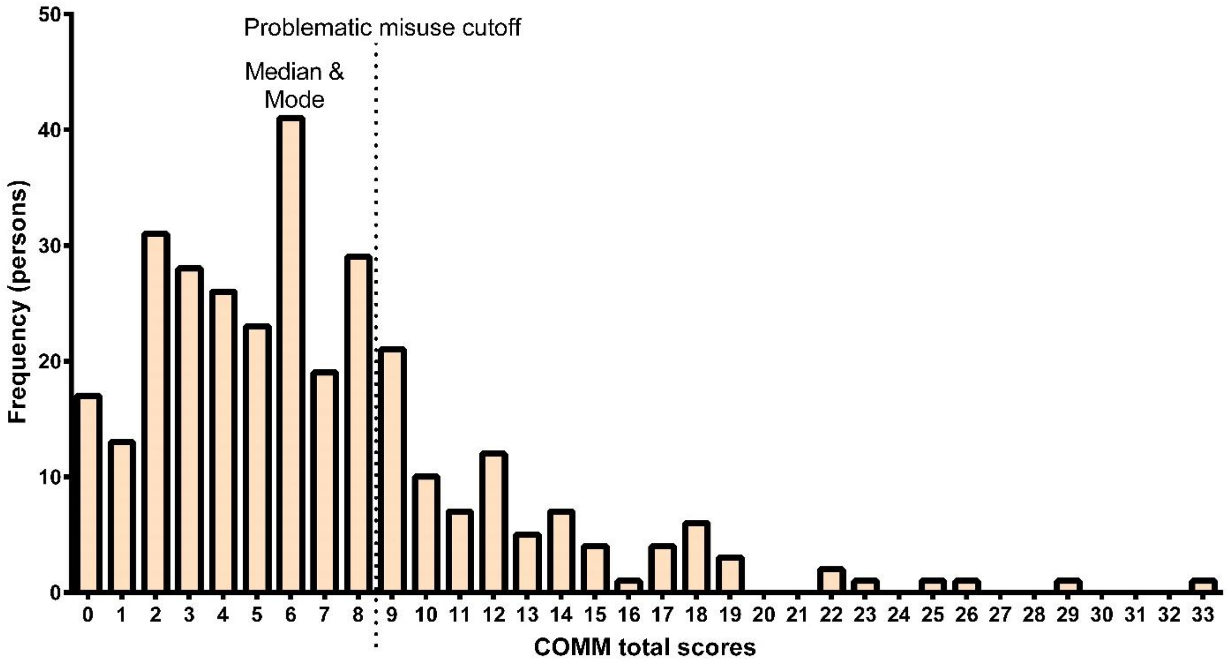

Figure S1. COMM score distribution

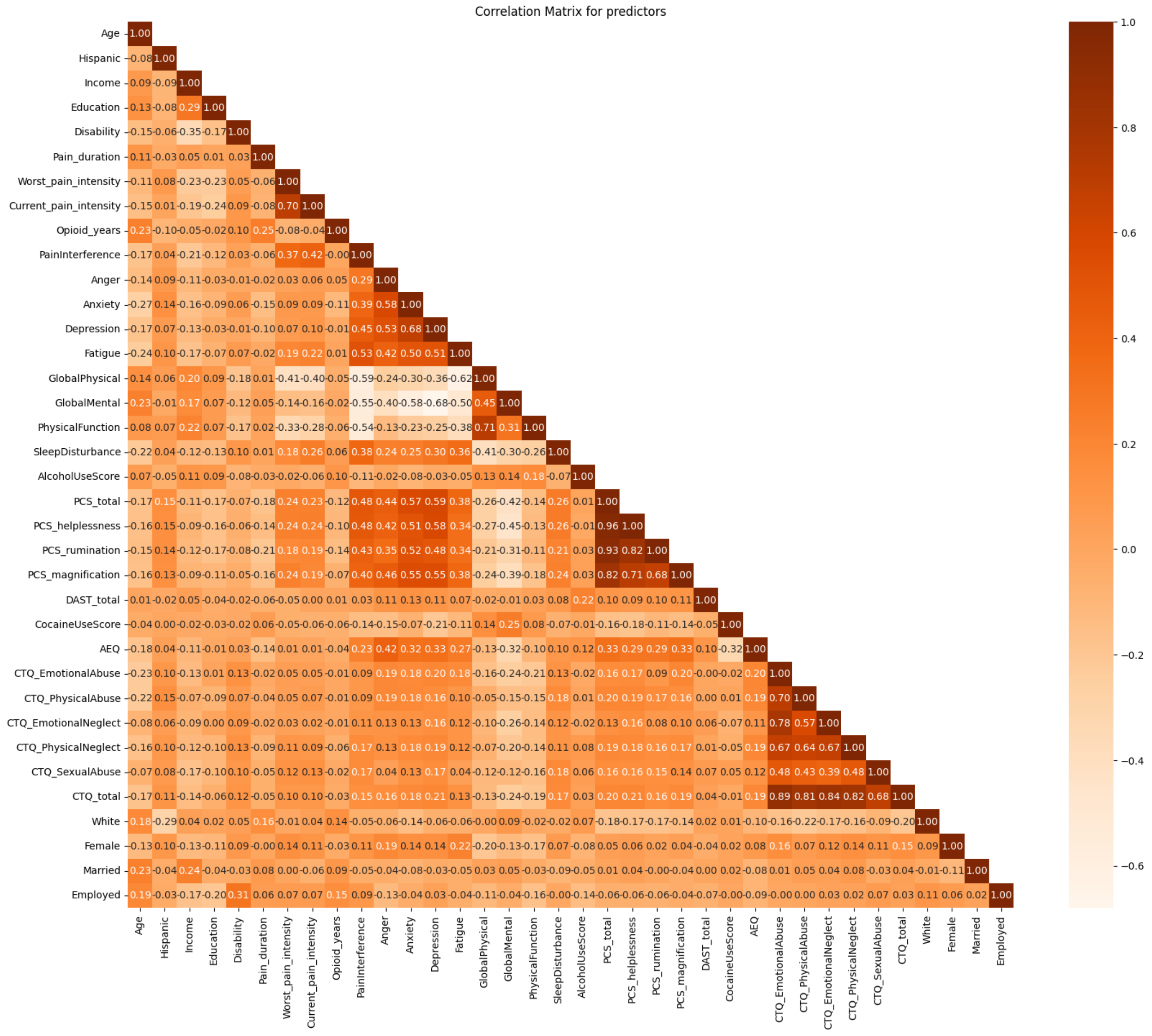


Figure S2. Correlation matrix between 36 predictors

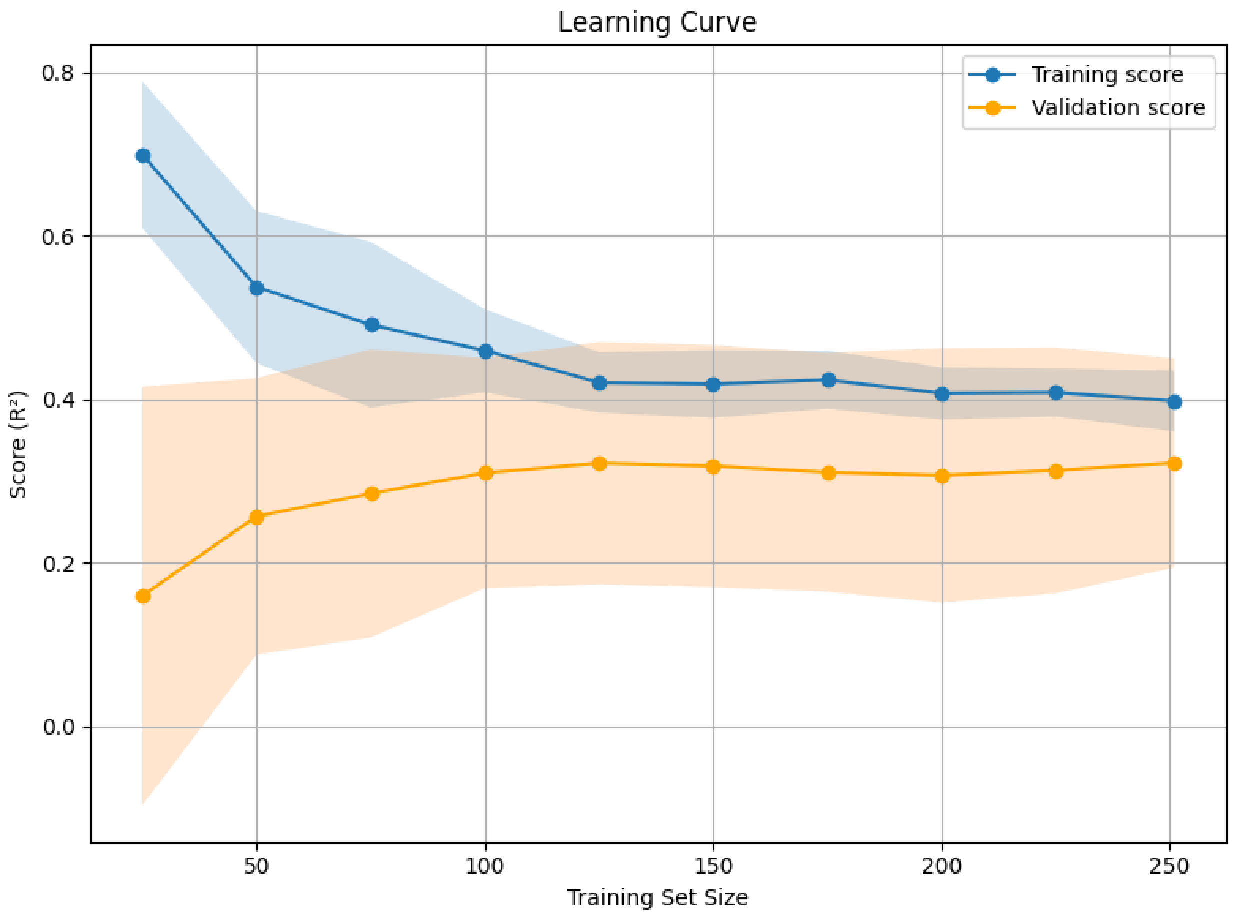


Figure S3. Learning curve analysis
